# Platelet-to-lymphocyte ratio (PLR), a novel biomarker to predict the severity of COVID-19 patients: a systematic review and meta-analysis

**DOI:** 10.1101/2020.08.21.20166355

**Authors:** Daniel Martin Simadibrata, Bashar Adi Wahyu Pandhita, Muammar Emir Ananta, Tamara Tango

## Abstract

**Background:** Platelet-to-lymphocyte ratio (PLR), a novel inflammatory marker, has been suggested to be able to predict the severity of COVID-19 patients. This systematic review aims to evaluate the association between PLR levels on admission and the severity of COVID-19 patients.

**Methods:** A systematic literature search was done on 23 July 2020 to identify peer-reviewed studies across four different databases (Ovid MEDLINE, EMBASE, SCOPUS, and the Cochrane Library), preprints from two databases (MedRxiv and SSRN), and grey literature from two databases (WHO COVID-19 Global Research Database and Center for Disease Control and Prevention COVID-19 Research Article). Research articles comparing the PLR value on admission in adult patients with COVID-19 with varying degrees of severity were included in the analysis. The following keywords were used for the search: “COVID-19”, “PLR”, “severity”, and “mortality”. The inverse variance method was used to calculate the pooled effect standardized mean difference (SMD) along with its 95% confidence interval (CI).

**Results:** A total of seven studies were included in the meta-analysis, six of which were conducted in China. From a total of 998 participants included, 316 (31.7%) had severe diseases; and those in the severe group were generally older and had underlying diseases compared to the non-severe group. In comparison to non-severe patients, the meta-analysis showed that severe COVID-19 patients had higher PLR levels on admission (SMD 0.68; 95%CI 0.43-0.93; I^2^ =58%).

**Conclusion:** High PLR levels on admission were associated with severe COVID-19 cases. Therefore, on-admission PLR level is a novel, cost-effective, and readily available biomarker with a promising prognostic role for determining the severity of COVID-19 patients.

## Introduction

Coronavirus Disease 2019 (COVID-19) is a disease caused by the severe acute respiratory syndrome coronavirus-2 (SARS-CoV-2), a virus thought to start as a zoonotic infection in Wuhan in late December 2019 (1). The disease was declared by the World Health Organization (WHO) as a pandemic on 11 March 2020 and has infected more than 100 countries worldwide. As of 26 July 2020, a total number of 15 785 641 cases and 640 016 deaths attributed to COVID-19 were recorded, only months after its emergence (2).

COVID-19 is known for being infectious and simultaneously manifesting in different organs aside from the pulmonary system (3–5). Patients infected with COVID-19 present a wide range of clinical conditions – ranging from asymptomatic infection, minimal symptoms to fatal respiratory distress. Although the majority of COVID-19 cases were classified as mild, involving flu-like symptoms to mild pneumonia, up to 20% of mild/moderate cases progressed to acute respiratory distress syndrome (ARDS) (6). Additionally, patients with rather normal clinical conditions can rapidly deteriorate and worsen within the course of a few days, making clinical presentation an unreliable prognostic predictor of COVID-19. Thus, a more objective indicator is required to accurately assess and stratify the prognosis of COVID-19 patients upon admission to healthcare services.

Immunological studies have shown that high levels of proinflammatory cytokines, known as cytokine storm, are the hallmark characteristic of severe COVID-19 cases. This extreme elevation of cytokines causes a massive proinflammatory response resulting in Multiple Organ Dysfunction Syndrome (MODS) and ARDS, which subsequently leads to mortality in COVID-19 patients (7). Therefore, in theory, inflammatory markers can be used to assess the severity and mortality risk of COVID-19 patients.

Platelet-to-lymphocyte ratio (PLR) is a novel marker of inflammation, which is inexpensive and readily available in clinical settings. PLR has been used in various diseases, such as cardiovascular diseases and autoimmune diseases, as a predictor of inflammation and mortality (8, 9). Due to the rapid involvement of inflammatory processes in COVID-19, severe COVID-19 patients have demonstrated elevated PLR levels on admission (10, 11). This suggests the potential of this inflammatory marker to determine the prognosis of COVID-19 patients, especially in resource-limited settings. Therefore, this systematic review aims to review the prognostic value of PLR levels on admission to determine the severity and mortality of COVID-19 patients.

## Methods

### Protocol and Registration

This systematic review was written in compliance to the Preferred Reporting Items for Systematic Reviews and Meta-analyses (PRISMA) Checklist **(Table S1)**. Prior to the writing of this systematic review, a protocol was formulated and registered in the International Prospective Register of Systematic Reviews (PROSPERO) on 2 June 2020 (CRD42020189369).

### Eligibility Criteria

We included cohort studies evaluating the difference in PLR levels on admission in adults (>18 years old) with confirmed COVID-19 (diagnosed using RT-PCR) categorized based on disease severity (severe and non-severe patients), and/or mortality (survivor and non-survivor). Case series, correspondences, review articles, non-research articles, and letters to editor were excluded from the study. We only included papers written and published in English. Otherwise, no other exclusion criteria were applied. The severity of the disease was defined by the WHO-China Joint Mission on COVID-19 report (12). Severe COVID-19 was defined as patients that met any of the following criteria: respiratory frequency ≥30x/minute, blood oxygen saturation ≤93%, PaO2/FiO2 ratio <300, and/or lung infiltrates >50% of the lung field within 24-48 hours.

### Search Strategy

The keywords used in the search strategy were derived from the following key concepts “COVID-19”, “platelet-to-lymphocyte”, “severity”, and “mortality”; and were adapted to the respective databases **(Table S2)**. A systematic literature search was finalized on 23 July 2020 to identify peer-reviewed papers published in four databases (Ovid MEDLINE, EMBASE, SCOPUS, and the Cochrane Library). Additionally, manual handsearching was done for preprints in two databases (MedRxiv and SSRN), and for grey literatures in two databases (WHO COVID-19 Global Research Database and Center for Disease Control and Prevention COVID-19 Research Article). We also performed forward and backward tracing of references from relevant articles to identify additional papers missed from database searching.

### Study Selection

All articles retrieved from the searches were exported to EndNote X9 reference manager. After de-duplication of articles, publications were screened based on its titles and abstracts, and the remaining publications were screened according to the full text. This study selection process was carried out by two independent reviewers (DMS and BAW). Any disagreement regarding the study selection was resolved by the inclusion of a third party.

### Quality Assessment and Data Extraction

All included articles’ data were extracted by two independent reviewers (DMS and BAW). Risk of bias assessment was done using the Newcastle Ottawa Scale (NOS). A standardized data extraction form was used to gather relevant data from the selected articles. Data extracted included: first author and publication year, publication type, country of study, study design, baseline characteristics of patients (age, gender, and underlying diseases), and the outcome of the study (PLR values).

### Statistical Analysis

For the quantitative analysis, we exported quantitative data from all eligible studies to Review Manager software 5.3 (Cochrane Collaboration) and performed a meta-analysis. We extrapolated the mean and standard deviation of studies with non-normal data using the available median and interquartile range (IQR) according to Hozo et al. (13). The inverse variance method was used to obtain the effect estimate in the form of standardized mean difference (SMD) and the 95% confidence Interval (CI). The statistical heterogeneity between the studies was assessed using Cochrane chi-square and I^2^. If there was significant heterogeneity (I^2^>50%), the random-effect model was used to calculate the pooled effect size, otherwise the fixed-effect model was used. Funnel plot analysis was used to qualitatively evaluate the risk of publication bias by comparing the SMD with the standard error of the natural log of SMD. We performed sensitivity analysis by omitting one study at a time to identify the source of heterogeneity. All p-values were two-tailed and statistical significance was considered if p<0.05.

## Results

### Search Selection and Study Characteristics

The literature search from electronic databases resulted in 27 publications, four of which were identified through manual handsearching of relevant literatures **(Figure 1; Figure S2)**. Following de-duplication, 17 articles were screened for its titles and abstracts. The remaining 10 articles were reviewed for its full-text and three articles were excluded, leaving a total of seven peer-reviewed articles included in this systematic review.

From a total of seven selected studies, all were peer-reviewed and were retrospective observational studies. In overall, six studies were done in China (10, 11, 14–17) and only one was done outside of China, in Turkey (18). All studies compared the on-admission PLR values between severe and non-severe COVID-19 patients, except for one that compared the PLR values between severe and moderate COVID-19 patients (17). Unfortunately, no study explored the prognostic value of PLR on admission to predict mortality. The quality of the included studies is shown in **Table S3**. Four studies had seven NOS stars and the other three had six NOS stars, in brief, all studies were of acceptable quality and eligible for inclusion in the meta-analysis.

**Figure 1.**
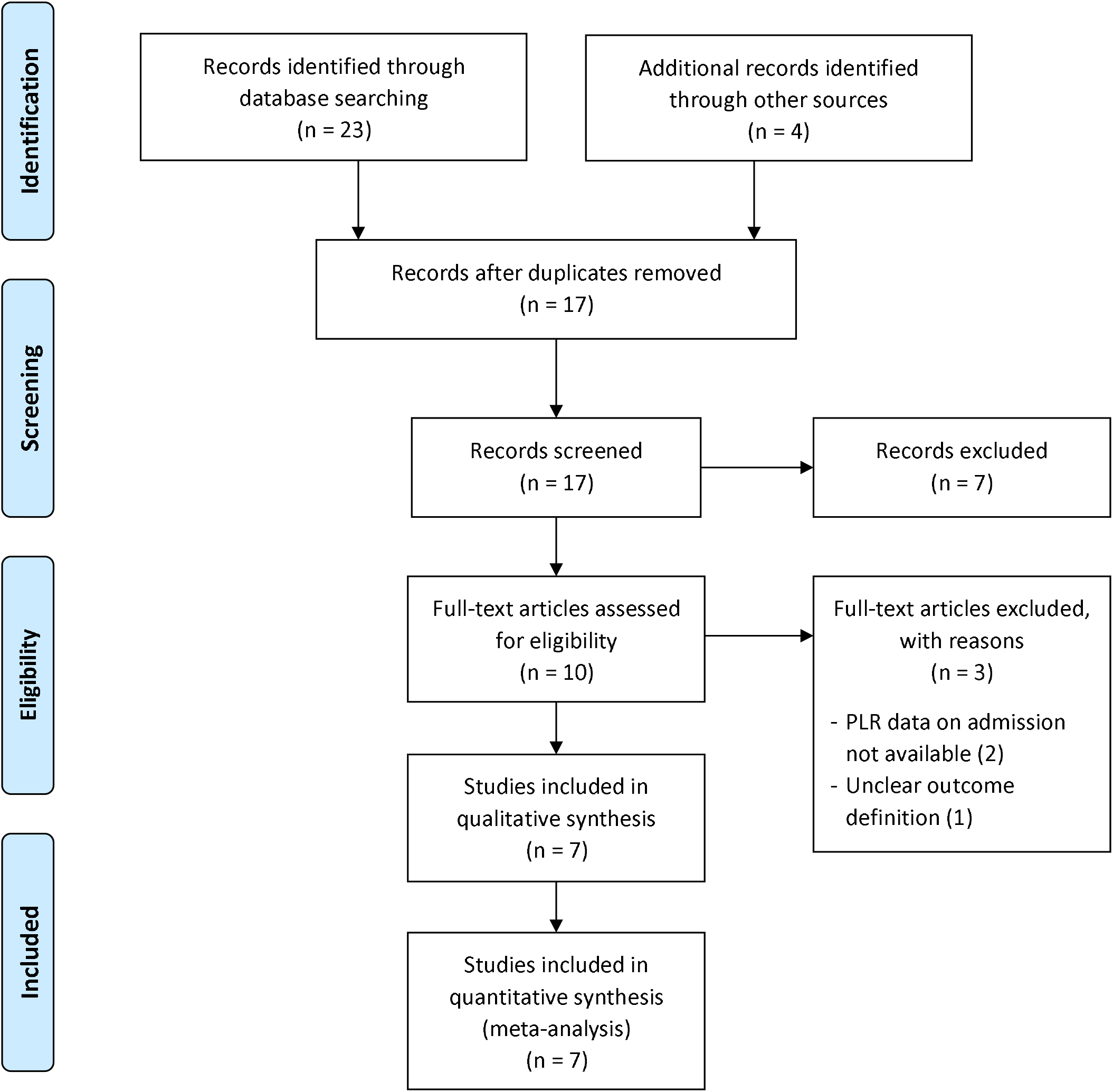
Prisma diagram for study selection in the meta-analysis. A systematic search of the literature was performed from database conception to 23 July 2020.

### Baseline Characteristics and Study Findings

A total of 998 participants were included from all the studies, 316 (31.7%) of which had a severe disease. Although four studies did not report the baseline comorbidity characteristics of COVID-19 patients, in comparison to non-severe COVID-19 patients, those with severe diseases were generally significantly older, and more likely to have underlying diseases such as hypertension, diabetes, and cardiovascular diseases. Of all the studies, only Qu et al. reported higher PLR values on admission in non-severe patients compared to severe patients (non-severe: 242.75 ± 173.74 vs severe: 160.02 ± 51.99; p=0.414) while the remainder demonstrated higher on-admission PLR values in severe compared to non-severe COVID-19 patients.

Two studies performed a receiver operator curve (ROC) analysis to determine the most optimal cut-off level for PLR. Yang et al. reported the optimal cut-off level for PLR was 180 [Area under the ROC Curve (AUC) 0.784 (95%CI 0.666-0.901); sensitivity 77%; specificity 44%] (11) while Sun et al. reported the optimal cut-off level was 226.67 [AUC 0.746 (95%CI 0.637-0.854); sensitivity 59.26% (95%CI 38.8%-77.6%); specificity 80.90% (95%CI 71.2%-88.5%)] (16).

### Platelet-to-lymphocyte Ratio (PLR) and Severity of COVID-19

A pooled effect size meta-analysis was conducted using the random-effect model (n=998; severe = 316, non-severe = 682) **(Figure 2)**. In overall, patients with severe COVID-19 had a higher PLR value on admission compared to non-severe COVID-19 (SMD 0.68; 95%CI 0.43 0. 93). There was a significant heterogeneity among the studies (I^2^=58%, p=0.03). The funnel plot was visually asymmetrical and indicated a potential risk of publication bias **(Figure S1)**.

**Figure 2.**
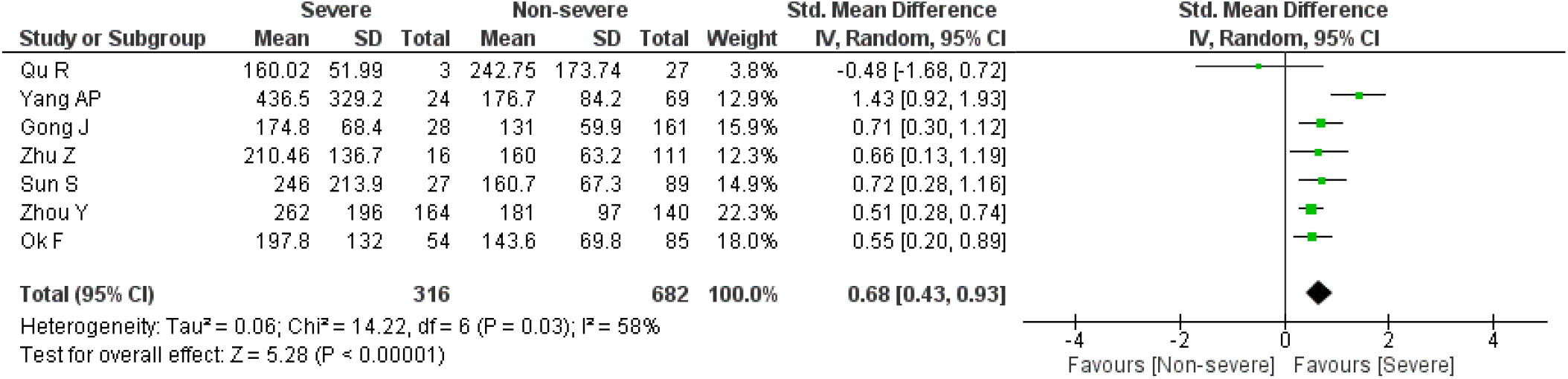
PLR value on admission and severity of COVID-19. Forrest Plot using the inverse variance random-effect model showing the association between PLR value on admission and severity of COVID-19 for all included studies.

Sensitivity analysis by sequentially removing one study at a time did not significantly change the heterogeneity among the studies and the overall pooled effect size. However, the exclusion of the study by Yang et al. resulted in no significant heterogeneity between the studies (I^2^=0%, p=0.52) **(Figure S2)**. The pooled effect size meta-analysis using the fixed-effect model showed a significantly higher PLR value on admission in severe COVID-19 patients compared to non-severe COVID-19 patients (SMD 0.57; 95%CI 0.41-0.72).

## Discussion

As of 26 July 2020, COVID-19 has infected approximately 15 million people worldwide, 600 thousand of whom died (2). More concerning, those with severe COVID-19 rapidly deteriorate to critical cases, which involve multiorgan failure, leading to death (19). Thus, there is an urgent need for healthcare providers to develop readily available biological markers to predict the severity and mortality of COVID-19 at the early stage of the disease to provide the most optimal management.

PLR has been initially suggested as a great candidate marker for determining the severity and mortality of COVID-19. First, PLR is an established marker of inflammation (20). Inflammation plays a considerable role in the pathophysiology of COVID-19, with cytokine storm as a hallmark condition in severe disease and poorer prognosis (21). Thus, elevated PLR value suggests an overactive inflammatory response and subsequently worse prognosis. Second, PLR is sensitive to natural and acquired immune response (22). Third, PLR is an inexpensive and readily available measurement that can be used in resource-limited settings. Therefore, our systematic review aims to review the validity of PLR level on admission as a prognostic indicator in COVID-19 patients.

Our meta-analysis, which included a total of 998 COVID-19 patients, showed that high PLR value was associated with severe COVID-19. Six out of the seven included studies demonstrated similar results with increased PLR on admission found in severe cases of COVID-19 compared to those with mild or moderate disease. This suggests that elevated PLR on admission among severe COVID-19 patients reflects a higher degree of cytokine storm. This evidence can be useful for the provision of specialized treatment to patients with severe COVID-19 as they might require longer hospital admissions.

Although PLR on admission was generally shown to be increased in severe vs non-severe COVID-19, Qu et al. reported a reduced PLR on admission in severe compared to non-severe COVID-19 despite no statistical significance. Inconsistencies between the results of different studies could be problematic, especially for daily clinical application. However, we noted the rather small sample size of the study, only three severe cases of COVID-19 were included in the analysis compared to 27 non-severe cases, which could lead to this contradictory result (10). Furthermore, follow-up analysis of PLR in the study showed elevated levels upon hospitalization.

To date, there is no universal laboratory reference value for PLR, especially for COVID-19 patients. Of all the included studies, only two studies attempted to determine the optimal cut-off PLR value. Yang et al. reported the optimal cut-off PLR value as 180 with AUC of 0.784, specificity of 44%, and sensitivity of 77% (11). Meanwhile, Sun et al. suggested a cut-off PLR value of 226.67 with AUC of 0.746, specificity of 80.90% and sensitivity of 59.26% (16). This discrepancy warrants the need for further research to determine the most appropriate PLR cutoff value in determining the severity of COVID-19 patients.

As PLR involves a comparison between the absolute platelet and absolute lymphocyte count, a comprehensive understanding about the role of platelets and lymphocytes in COVID-19 is important. Platelet is a marker for inflammation in various diseases (8, 9). The platelet level in severe COVID-19 varies between studies. From the seven studies, six studies showed low platelet count in severe compared to non-severe group, two of which were significantly different statistically. Three hypotheses may explain the underlying decrease in platelet count. First, the cytokine storm triggered by SARS-CoV-2 can decrease the synthesis of platelets, by destroying bone marrow progenitor cells. SARS-CoV-2 is also postulated to directly affect the production of platelets in the bone marrow. Second, SARS-CoV-2 can induce the generation of autoantibody and immune complex which may trigger the destruction of platelets. Third, platelets activated during lung injury could be aggregated and be overactively involved in microthrombus formation (23). However, there is still a lack of current evidence for platelet cut-off level in COVID-19.

Moreover, from the six studies showing increased PLR levels on admission in severe cases, the absolute lymphocyte count was also decreased. Another study also reported lymphopenia among severe COVID-19 (24). The underlying mechanism for this decreased absolute lymphocyte count is that SARS-CoV-2 triggers pyroptosis in lymphocytes through the activation of NLRP 3 inflammasome (23). Another hypothesis points out the role of the pro-inflammatory cytokine IL-6 that uses the lymphocytes, thus decreased lymphocyte counts are associated with poor prognosis. This evidence of lymphopenia has been reported from the meta-analysis by Huang et al., which showed lymphocyte count less than or equal to 1100 cells/μL was associated with threefold risk of poor outcome in severe COVID-19 cases (25).

We also observed a significant heterogeneity among the included studies in this meta-analysis. The possible reasons for the high heterogeneity could be due to the distinct interstudy baseline characteristics of the subjects, different number and proportion of patients with comorbidities as well as the proportion of severe and non-severe cases. Fesih, Qu, and Sun excluded patients with other comorbidities, such as chronic lung diseases, hematological diseases, and liver diseases (10, 16, 18) while Gong did not provide any data on the comorbidities of the patients (14). Another reason could be due to the small number of included studies in this meta-analysis, which could reduce the accuracy of the heterogeneity analysis (26). We performed sensitivity analysis by sequentially omitting one study at a time and determined that Yang et al. was the source of heterogeneity (11). However, with the removal of Yang et al., pooled analysis still showed a statistically significant higher PLR value on admission in severe COVID-19 in comparison to non-severe COVID-19 patients.

This meta-analysis is not without limitations. We acknowledge that only including articles written and published in English would disregard those written in other languages, and thus present with geographical bias. Moreover, most of the included studies were from China, whereas the majority of confirmed cases and deaths were located in the USA and Europe. The variability in PLR values between different populations could limit the relevance of this finding. Based on the funnel plot, we also identified a potential risk of publication bias. In addition, the limited data presented by the included studies did not allow further stratification of the severe group into severe and critically ill patients. Therefore, further research still needs to be conducted to determine an optimal cut-off value for PLR value for the prediction of severity in COVID-19.

## Conclusion

Our meta-analysis showed that PLR can be used as a novel, cost-effective, and readily available biomarker in determining the severity of COVID-19 patients. The implication of our finding is that elevated PLR levels on admission can be utilized as a prognostic indicator of severity in COVID-19 patients, especially in resource-limited settings, where there is an urgent need to effectively allocate medical resources and divert attention to patients with poorer prognosis. However, further studies are needed to determine the cutoff value of PLR with the most optimal sensitivity and specificity prior to adaptation in clinical practice.

## Data Availability

All data generated or analyzed during this study are included in the published article and its supplementary information files. The corresponding author (DMS) can be contacted for more information.

**Abbreviations**
ARDS: Acute respiratory distress syndrome;
AUC: Area under the ROC curve;
CI: Confidence interval;
COVID-19: Coronavirus disease 2019;
IQR: Interquartile range;
MODS: Multiple organ dysfunction syndrome;
NOS: Newcastle Ottawa Scale;
PLR: Platelet-to-lymphocyte ratio;
PRISMA: Preferred reporting items for systematic reviews and meta-analyses;
PROSPERO: International prospective register of systematic reviews;
ROC: Receiver operator curve;
SARS-CoV-2: Severe acute respiratory syndrome coronavirus-2;
SD: Standard deviation;
SMD: Standardized mean difference;
WHO: World Health Organization

## Declarations

### Ethics and approval and consent to participate

Not applicable.

### Consent for publication

Not applicable.

### Authors’ Contributions

**DMS and BAWP**: Idea formulation, article draft writing, data collection and analysis, interpretation of the data, critical review of the writing; **TT**: Article draft writing, data collection, interpretation of the data, critical review of the writing; **MEA**: Article draft writing, critical review of the writing. All authors contributed substantially to the writing of this manuscript and have consented for this submission.

## Acknowledgements

None.

---

**Table 1.** Summary of baseline characteristics and study findings of all included studies in the meta-analysis

| Io | Author | Study Design | Groups | Sample (N) | Male/Female (% of male) | Age (years) Mean ± SD / Median (IQR) | *p* | HT N (%) | *p* | DM N (%) | *p* | CVD N (%) | *p* | PLR value Mean ± SD / Median (IQR) | *p* |
| --- | --- | --- | --- | --- | --- | --- | --- | --- | --- | --- | --- | --- | --- | --- | --- |
| | Qu R | Retrospective Observational | **Non-severe** | 27 | 16/14 (53) | 49.4 ± 14.9 | 0.041* | NR | NR | NR | NR | NR | NR | 242.75 ± 173.74 | 0.414 |
| | | | **Severe** | 3 | | 60.0 ± 5.3 | | NR | | NR | | NR | | 160.02 ± 51.99 | |
| | Yang AP | Retrospective Observational | **Non-severe** | 69 | 38/31 (55) | 42.1 ± 18.6 | 0.034* | 7 (10) | <0.01* | 8 (12) | <0.01* | 4 (6) | <0.01* | 176.7 ± 84.2 | <0.001* |
| | | | **Severe** | 24 | 18/6 (75) | 57.9 ± 11.8 | | 16 (67) | | 13 (54) | | 9 (38) | | 436.5 ± 329.2 | |
|  | Gong J | Retrospective Observational | **Non-severe** | 161 | 72/89 (45) | 45.0 (33.0-62.0) | <0.01* | NR | NR | NR | NR | NR | NR | 131.0 (96.6-177.4) | 0.05 |
|  |  |  | **Severe** | 28 | 16/12 (57) | 63.5 (54.5-72.0) |  | NR |  | NR |  | NR |  | 174.8 (117.7-210.0) |  |
| | Zhu Z | Retrospective Observational | **Non-severe** | 111 | 73/38 (66) | 49.9 ± 15.5 | 0.03* | 23 (21) | 0.025* | 10 (9) | 0.451 | 4 (4) | 0.348 | 160.00 (129.64-215.00) | 0.299 |
| | | | **Severe** | 16 | 9/7 (56) | 57.5 ± 11.7 | | 8 (50) | | 0 (0) | | 2 (13) | | 210.46 (116.33-300.88) | |
|  | Sun S | Retrospective Observational | **Common** | 89 | 42/47 (47) | 47.0 (37.0-54.5) | <0.001* | NR | NR | NR | NR | NR | NR | 160.7 (116.7-207.5) | <0.001* |
|  |  |  | **Severe** | 27 | 18/9 (67) | 62.0 (53.0-71.0) |  | NR |  | NR |  | NR |  | 246.0 (167.9-456.7) |  |
| | Zhou Y | Retrospective Observational | **Moderate** | 140 | 55/85 (39) | 55.9 ± 14.4 | NR | NR | NR | NR | NR | NR | NR | 181 ± 97 | NR |
| | | | **Severe** | 123 | 58/65 (47) | 63.8 ± 13.9 | | NR | | NR | | NR | | 262 ± 196 | |
| | | | **Critically severe** | 41 | 25/16 (61) | 65.2 ± 12.7 | | NR | | NR | | NR | | | |
| | Ok F | Retrospective Observational | **Non-severe** | 85 | 38/47 (45) | 47.2 ± 15.7 | <0.001* | 9 (11) | <0.001* | 12 (14) | 0.218 | 2 (2) | <0.001* | 143.6 (94.2) | 0.072 |
| | | | **Severe** | 54 | 24/30 (44) | 68.3 ± 14.9 | | 24 (44) | | 12 (22) | | 17 (32) | | 197.8 (178.2) | |

**Notes**
CVD = Cardiovascular Disease
DM = Diabetes Mellitus
HT = Hypertension
QR = Interquartile Range
NR = Not reported
*p* = *p*-value; \* = statistically significant (*p*<0.05)
PLR = Platelet-to-lymphocyte ratio
SD = Standard Deviation

